# Genomic Epidemiology and Genetic Relatedness of Antimicrobial-Resistant *Morganella morganii* in Community and Hospital Settings in Bangladesh

**DOI:** 10.64898/2026.09.23.26363554

**Authors:** Tanzim Rahman, Tahsin Khan, Arefeen Haider, Shovan Basak Moon, Nure Sharaf Nawar Samia, Syeda Mah-E-Muneer, Gazi Md Salahuddin Mamun, Fahmida Chowdhury, Mustafizur Rahman, Mohammad Jubair

## Abstract

*Morganella morganii* is an opportunistic pathogen with intrinsic and acquired antimicrobial resistance, but its genomic epidemiology remains poorly characterized in Bangladesh. We sequenced and compared genomes of 47 stool-derived *M. morganii* isolates from hospital (n = 29) and community (n = 18) participants in urban Dhaka.

Susceptibility testing showed frequent resistance to imipenem (70·2%) and cefuroxime (97·9%), while all isolates were resistant to colistin. Resistance to meropenem (6·4%) and ertapenem (2·1%) was uncommon. Based on acquired non-susceptibility across tested antimicrobial categories, 18/47 (38·3%) isolates were multidrug-resistant, including 15/29 (51·7%) hospital-derived and 3/18 (16·7%) community-derived isolates. Genomic analysis identified 17 β-lactamase alleles including *Morganella*-associated chromosomal β-lactamases and acquired resistance determinants, with *bla*_OXA-1_, *bla*_DHA-21_, *bla*_CTX-M-15_, and *bla*_TEM-1_ predominating. However, no recognized acquired carbapenemase genes were detected. Among 44 non-β-lactam resistance genes, *tetB* was most frequent.

Phylogenomic and average nucleotide identity analyses classified 31 isolates as *M. morganii* subsp. *morganii* and 16 as the subsp. *intermedius* lineage. Considerable population diversity was observed, with 33 distinct sequence types among the 47 isolates and no clear clustering by hospital or community origin. The subsp. *intermedius* isolates showed greater genomic heterogeneity than subsp. *morganii*, with a minimum within-group average nucleotide identity of 94·2%. Single nucleotide variant (SNV) analysis identified several closely related isolate groups, including an ST1 pair differing by 10 SNVs and collected three days apart within the same hospital, although direct transmission could not be established.

Virulence-associated gene profiles differed between subspecies, with hemolysin- and toxin-associated genes predominantly detected among subsp. *morganii* isolates and largely absent from subsp. *intermedius*. Overall, these findings reveal substantial genomic diversity, a considerable burden of multidrug resistance, and subspecies-associated differences in virulence gene content among *M. morganii* circulating in Dhaka, while the frequent imipenem-non-susceptible but carbapenemase-negative phenotype highlights the need to investigate alternative carbapenem resistance mechanisms.

## INTRODUCTION

*Morganella morganii* is a Gram-negative, facultative anaerobic bacterium belonging to the order Enterobacterales and the family Morganellaceae. Although it commonly colonizes the human gastrointestinal tract as part of the normal gut microbiota, it is increasingly recognized as an opportunistic pathogen associated with healthcare-associated infections worldwide.^1–3^ *M. morganii* exhibits intrinsic resistance to several antimicrobials, including aminopenicillins, most first- and second-generation cephalosporins, colistin, and macrolides, through multiple mechanisms. Its chromosomally encoded inducible AmpC β-lactamase contributes specifically to intrinsic β-lactam resistance.^2,4,5^ Consequently, infections caused by *M. morganii* can be difficult to treat and have been associated with increased morbidity and mortality owing to delays in appropriate empirical antimicrobial therapy.^6^ The emergence of carbapenem resistance in *M. morganii* is therefore clinically concerning, particularly as carbapenem-resistant Enterobacterales are classified as critical-priority pathogens in the 2024 WHO Bacterial Priority Pathogens List.^7^ However, carbapenem resistance in *M. morganii* may present with discordant susceptibility across individual carbapenems, including resistance to imipenem despite retained susceptibility to meropenem and ertapenem, complicating interpretation within the broader CRE framework.^8^

Carbapenem resistance in *M. morganii* has been associated with acquisition of carbapenemase genes, including *bla*_NDM-1_, *bla*_KPC-2_, *bla*_IMP-like_, and *bla*_OXA-48_, which are among the most frequently reported carbapenemase determinants in global *M. morganii* genomes while resistance to extended-spectrum cephalosporins is commonly mediated by *bla*_DHA-like_ and *bla*_MOR-2_ genes.^9,10^ Beyond antimicrobial resistance (AMR), the pathogenic potential of *M. morganii* is influenced by a range of virulence-associated genes involved in host colonization, immune evasion, iron acquisition, and toxin production.^11^ However, the distribution of these virulence determinants across the different *M. morganii* lineages remains poorly characterized. Genomic epidemiology plays an integral role in understanding pathogenic bacteria, offering insights into their transmission routes, virulence, drug resistance, genetic variations, and evolutionary trends.

Recent whole-genome sequencing studies have substantially improved our understanding of the population structure and antimicrobial resistance of *M. morganii*. Li et al.^9^ analyzed the phylogenomics of 968 *Morganella* spp. isolates collected from 52 countries, with the majority originating from China and the United States. Additional genomic investigations have characterized the diversity, resistance mechanisms, and epidemiology of *M. morganii* isolates in Europe and China.^5,12,13^ However, comparable genomic studies from South Asia, particularly Bangladesh, remain lacking.

Bangladesh is recognized as a hotspot for antimicrobial resistance, with high levels of resistance among Enterobacterales isolated from both clinical infections and asymptomatic carriage, reflecting extensive antimicrobial use and limitations in antimicrobial stewardship and surveillance.^14,15^ As *M. morganii* commonly colonizes the intestinal tract, it may serve as an important reservoir for resistance determinants that can persist and disseminate between community and hospital settings. Characterizing the genomic diversity, antimicrobial resistance, and virulence potential of *M. morganii* circulating in Bangladesh is therefore essential to better understand its contribution to the regional AMR burden.

The taxonomy of *Morganella morganii* has undergone substantial revision and remains an area of active investigation. Traditionally, *M. morganii* was divided into two subspecies, subsp. *morganii* and subsp. *sibonii*, based largely on phenotypic characteristics and DNA-DNA hybridization.^16,17^ More recent whole-genome analyses have revealed greater genomic diversity within *Morganella*. Bonnin et al.^12^ proposed a revised genomic framework distinguishing the *sibonii* lineage from *M. morganii* and recognizing a proposed *M. morganii* subsp. *intermedius* lineage alongside subsp. *morganii*. These findings highlight the value of genome-based approaches for resolving the population structure and taxonomic relationships of *M. morganii*.

In this study, we utilized whole-genome sequencing together with phenotypic antimicrobial susceptibility profiles to characterize the stool-derived *M. morganii* isolates collected from community and hospital settings in urban Dhaka, Bangladesh. We investigated their phylogenetic relationships, subspecies distribution, sequence type diversity, antimicrobial resistance genes, and virulence-associated genes to characterize the population structure and genomic epidemiology of *M. morganii* circulating in Bangladesh. In addition, single nucleotide variant (SNV) analysis was performed to investigate the genetic relatedness of closely related isolates and identify potentially epidemiologically linked isolates between community and healthcare settings.

## METHODS

### Participant enrollment

This study was conducted as part of the Antibiotic Resistance in Communities and Hospitals (ARCH) project, a multicountry study carried out across six countries to estimate the population-level prevalence of colonization with clinically significant antimicrobial-resistant organisms in community and hospital settings. Participants aged ≥18 years were enrolled between April and October 2019 from Kamalapur, an established urban community surveillance site in Dhaka, Bangladesh, and three participating hospitals (one tertiary-level government hospital and two private hospitals) that primarily served this community population. Hospital participants were recruited from adult inpatient departments, excluding paediatric and postoperative wards, using probability proportional to patient population size sampling. Healthy community participants were recruited from the hospital catchment communities within the Kamalapur surveillance area using a two-stage cluster sampling strategy. The study protocol was reviewed and approved by the Research Review Committee and Ethical Review Committee of icddr,b. Written informed consent was obtained from all participants in accordance with the approved study protocol. All patient IDs were de-identified to isolate IDs which were not known to anyone outside the research group, and only deidentified isolate IDs are provided in the manuscript.

### Microbiological Screening and Isolate Processing

Stool samples were collected from 1,433 participants in total recruited from hospitals (n = 719) and surrounding communities (n = 714). Samples were cultured on CHROMagar ESBL, CHROMagar mSuperCARBA, and CHROMagar COL-APSE selective media at the Clinical Microbiology Laboratory of icddr,b to recover antimicrobial-resistant Enterobacterales. Isolates were identified and subjected to antimicrobial susceptibility testing (AST) using the bioMérieux VITEK 2 system according to the Clinical and Laboratory Standards Institute (CLSI) 2019 guidelines.^18^ Multidrug resistance (MDR) was classified according to the definitions proposed by Magiorakos et al.^19^ MDR was defined as acquired non-susceptibility to at least one agent in three or more antimicrobial categories, with intermediate and resistant phenotypes considered non-susceptible. Four acquired-resistance categories represented in the phenotypic testing panel were considered: fluoroquinolones (ciprofloxacin), extended-spectrum cephalosporins (ceftriaxone, cefepime, and cefoperazone/sulbactam), carbapenems (ertapenem and meropenem), and folate pathway inhibitors (trimethoprim/sulfamethoxazole). Colistin and cefuroxime were excluded from MDR classification because resistance to polymyxins and first- and second-generation cephalosporins is intrinsic to *M. morganii*.^5,12,20^ Imipenem was also excluded from MDR classification because *Morganella* spp. exhibit intrinsic low-level resistance to imipenem due to reduced affinity of their penicillin-binding proteins, independent of carbapenemase production.^21^ Differences in the proportion of MDR isolates between hospital- and community-derived isolates and between subsp. *morganii* and subsp. *intermedius* were assessed using two-sided Fisher’s exact tests because of the relatively small group sizes. A p-value <0·05 was considered statistically significant. Extensive drug resistance (XDR) and pandrug resistance (PDR) classifications were not assigned because the antimicrobial susceptibility panel did not encompass all or nearly all antimicrobial categories required for these definitions.

The primary multi-country ARCH protocol included screening for extended-spectrum cephalosporin-resistant Enterobacterales (ESCrE) and carbapenem-resistant Enterobacterales (CRE) from stool samples. However, in the Bangladesh site, screening for colistin-resistant Enterobacterales (ColRE) was additionally incorporated. Enterobacterales isolates recovered through this screening were subsequently transferred to the icddr,b Genome Centre for whole-genome sequencing.

### DNA Extraction, Library Preparation, and Sequencing

Single colonies grown on MacConkey agar were subcultured and incubated for 24 hours before inoculation into Mueller-Hinton broth to obtain sufficient bacterial biomass for DNA extraction. Genomic DNA was extracted using the DNeasy Blood & Tissue Kit (Qiagen, Hilden, Germany) according to the manufacturer’s protocol. DNA libraries were prepared using the Illumina DNA Prep Kit (Illumina Inc., San Diego, CA, USA), with automated liquid handling performed on the epMotion 5075 platform (Eppendorf, Hamburg, Germany). Whole-genome sequencing was performed on the Illumina NextSeq 550 platform using 2 × 150 bp paired-end chemistry.

### Bioinformatics Analysis Workflow

Whole-genome sequencing data were processed using the PhoeNIx pipeline v2.1.1^22^ with default parameters. In brief, PhiX174 control sequences and adapter contamination were removed using BBDuK v39.01 (https://github.com/BioInfoTools/BBMap), followed by quality filtering and trimming with fastp v0.23.4.^23^ Trimmed reads were assembled *de novo* using SPAdes v3.15.5^24^ and contigs shorter than 500 bp were excluded from downstream analyses. Genome annotation was performed using Prokka v1.14.5.^25^ Taxonomic classification was performed by comparing assembled genomes against the NCBI RefSeq bacterial database using Mash v2.3^26^ and FastANI v1.33,^27^ with Kraken2 v2.1.3,^28^ using the Standard-8 database (July 2025 collection) (https://benlangmead.github.io/aws-indexes/k2), providing an additional quality control measure to assess contamination and an alternative taxonomic assignment where required.

Isolates identified as *M. morganii* by the PhoeNIx taxonomic workflow were retained for subsequent genomic analyses in this study.

### Subspecies Assignment, Phylogenomics, and Sequence Typing

To assign isolates to genomic lineages, 15 publicly available *Morganella* genomes representing subsp. *morganii*, the proposed subsp. *intermedius* lineage, and the *sibonii* lineage were selected from Bonnin et al.^12^ Five reference genomes from each subspecies were included together with an additional *M. morganii* reference genome (NCBI GenBank accession GCA_006094455.1). The complete list of reference genomes is provided in Supplementary Table S1.

A pangenome analysis of all *M. morganii* identified study isolates and the 16 reference genomes was performed using Panaroo v1.5.2,^29^ retaining genes present in at least 98% of genomes to generate a core genome alignment. SNP-sites v2.5.1^30^ was used to extract variable sites and count invariant sites from the alignment. These data were subsequently used to infer a maximum-likelihood phylogenetic tree with IQ-TREE v3.0.1^31^ under the GTR+F substitution model with 1,000 ultrafast bootstrap replicates. The resulting tree was visualized and annotated using iTOL.^32^

To further assess genomic relatedness and support subspecies assignment, pairwise average nucleotide identity (ANI) values were calculated between all study and reference genomes using pyANI v0.2.12^33^ using the ANIm method. Pairwise ANI values were interpreted with reference to the approximately 95% ANI threshold commonly used for prokaryotic species delineation, as supported by the large-scale genomic analysis of Jain et al.^27^ ANI values were summarized across all subsp. *morganii* and subsp. *intermedius* genomes combined and separately within each subspecies. Sequence types (STs) were assigned from assembled genome sequences using the BIGSdb-Pasteur platform (https://bigsdb.pasteur.fr/).

### Detection of Antimicrobial Resistance Genes

Antimicrobial resistance genes were identified using GAMMA v2.2^34^ within the PhoeNIx pipeline, which integrates NCBI AMRFinderPlus v3.12.8^35^ and the ARG-ANNOT^36^ and ResFinder^37^ databases. GAMMA-identified genes are filtered to only include those with ≥98% AA identity and ≥90% gene length.

### Virulence-Associated Gene Detection

Because *Morganella*-specific virulence factors were not included in the PhoeNIx virulence module at the time of analysis, a custom virulence screening approach was implemented. A curated list of *Morganella*-associated virulence genes was compiled from Chen et al.^11^ and is provided in Supplementary Table S6. These sequences were used as queries in BLAST v2.17.0^38^ searches against the assembled genomes. Hits were considered positive if they met a minimum percentage identity of 90% and query sequence coverage of 70%.

### Single Nucleotide Variant (SNV) Analysis

To investigate genomic relatedness and identify isolates with potential epidemiological links, pairwise SNV distances were calculated using the PhyloPhoeNIx v1.0 pipeline (https://github.com/CDCgov/phylophoenix), which utilizes SNVPhyl v1.8.2^39^ for SNV identification and distance matrix generation.

An initial SNV analysis was performed separately for the *M. morganii* subsp. *morganii* and *intermedius* groups to check the SNVPhyl core estimate. Due to relatively low SNVPhyl core genome coverage estimates (<90%), isolates were subsequently stratified by sequence type and SNV analyses were repeated for all STs represented by multiple isolates. This stratification increased the proportion of genomic positions included in the core alignment and enabled comparisons among more closely related isolates.

A validated SNV threshold for inferring recent transmission has not been established for *M. morganii*, although Bonnin et al.^12^ used a core-genome SNP (cgSNP) distance of <20 SNPs to identify closely related isolates potentially associated with recent transmission. In the absence of a validated transmission threshold for *M. morganii*, pairs differing by <100 SNVs were selected as a broad screening set for detailed comparison of temporal and epidemiological metadata.

### Data Visualization

All downstream data processing and analysis were performed using the pandas v2.2.3^40,41^ library. Data visualization was performed using Matplotlib v3.10.0^42,43^ and Seaborn v0.13.2.^44^

## RESULTS

### Isolate Selection and Characteristics

Following whole-genome sequencing and taxonomic classification using the PhoeNIx pipeline, 47 isolates were identified as *Morganella morganii* and included in the present study. Of these, 29 (61·7%) were obtained from hospital participants and 18 (38·3%) from community participants. Prior to whole-genome sequencing, species identification using the VITEK 2 system classified 41 isolates as *M. morganii* subsp. *morganii*, two as *M. morganii* subsp. *sibonii*, and four as *E. coli*. According to their original ARCH screening classifications, 41 (87·2%) isolates were categorized as CRE, four (8·5%) as ESCrE, and two (4·3%) as ColRE. Among the ESCrE isolates, two had been identified by VITEK 2 as *M. morganii* subsp. *morganii* and two as *E. coli*, while both ColRE isolates had initially been identified as *E. coli*. Thus, six isolates ultimately identified as *M. morganii* by WGS had received a different species or subspecies assignment by VITEK 2. Genome assembly and quality statistics for all 47 isolates are provided in Supplementary Table S3.

### Subspecies-Level Assignment

Subspecies-level classification, determined using both pangenome-based phylogenetic analysis and ANI comparisons with reference genomes representing the *M. morganii* subsp. *morganii*, *intermedius*, and *sibonii* lineages, is shown in Figure 1. Of the 47 isolates, 31 (66·0%) clustered with the subsp. *morganii* reference genomes and 16 (34·0%) with the proposed subsp. *intermedius* reference genomes. No study isolates clustered with the *sibonii* reference lineage.

**Figure 1:**
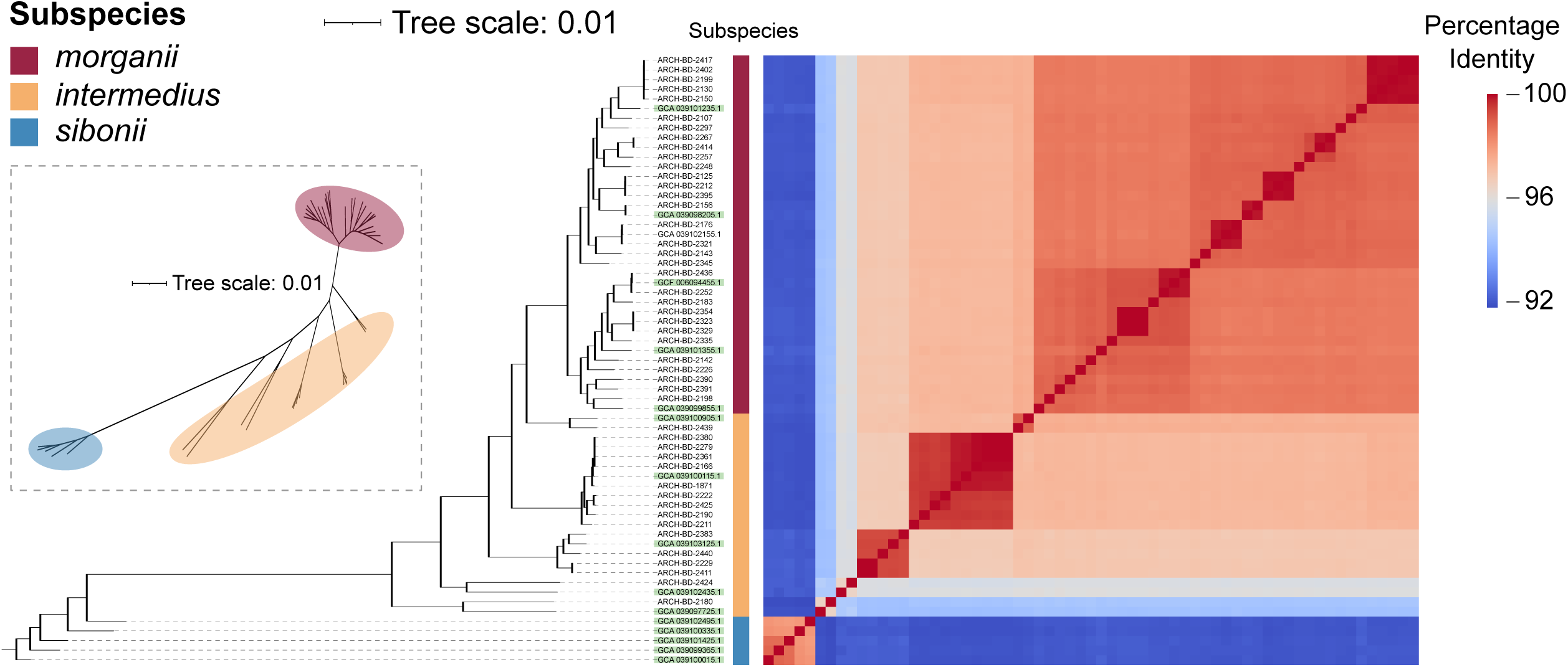
Maximum-likelihood phylogenetic tree and pairwise ANI matrix of the 47 study isolates together with reference genomes representing *M. morganii* subsp. *morganii*, subsp. *intermedius*, and subsp. *sibonii*. Subspecies assignments are indicated by annotation alongside the tree. Reference genome sample IDs are highlighted with a green background. The inset shows an unrooted phylogenetic tree illustrating the separation of the three genomic lineages.

Pairwise ANI analysis further demonstrated differences in genomic diversity between the two lineages. Across subsp. *morganii* and subsp. *intermedius* genomes combined, pairwise ANI values had a minimum of 94·2% and a median of 97·4%. Subsp. *morganii* showed comparatively high within-lineage similarity, with a minimum ANI of 98·4% and a median of 98·7%. In contrast, subsp. *intermedius* was more heterogeneous, with a minimum ANI of 94·2% and a median of 96·7%. Notably, two subsp. *intermedius* genomes (one isolate from the present study and one reference genome from Bonnin et al.^12^) showed <95% ANI with all other subsp. *morganii* and subsp. *intermedius* genomes, while retaining an ANI of 96·4% with each other.

### Antimicrobial Susceptibility Profiles

Antimicrobial susceptibility testing was performed against imipenem, ertapenem, meropenem, cefuroxime, cefoperazone/sulbactam, ceftriaxone, cefepime, ciprofloxacin, trimethoprim/sulfamethoxazole, and colistin. The distribution of minimum inhibitory concentrations (MICs) and susceptibility classifications across the 47 *M. morganii* isolates is presented in Figure 2. Resistance varied considerably among the three carbapenems tested. Resistance to imipenem was observed in 33 (70·2%) isolates, with a further 12 (25·5%) classified as intermediate and two (4·3%) as susceptible. In contrast, resistance to ertapenem and meropenem was detected in only 1 (2·1%) and 3 (6·4%) isolates, respectively, with the majority remaining susceptible to both antibiotics.

**Figure 2:**
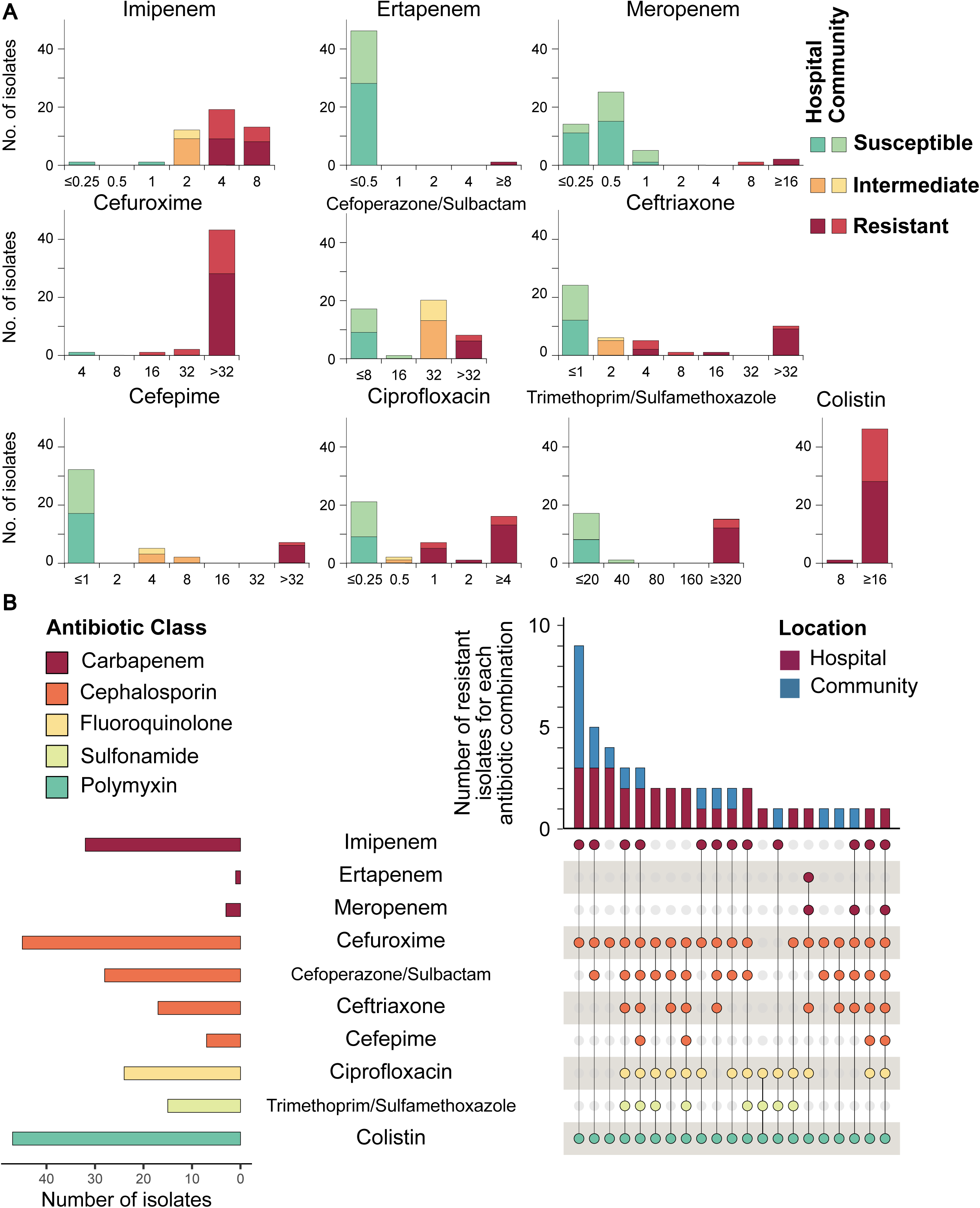
(A) Distribution of minimum inhibitory concentrations (MICs) for the tested antimicrobial agents among the 47 *M. morganii* isolates. Bars indicate the number of isolates at each MIC and are stratified by collection source (hospital or community) and antimicrobial susceptibility category (susceptible, intermediate, or resistant). (B) UpSet plot showing the frequency and combinations of antimicrobial resistance phenotypes across isolates. Horizontal bars indicate the total number of isolates resistant to each antimicrobial, while vertical bars represent the number of isolates exhibiting each resistance combination, stratified by hospital and community origin. Antimicrobials are grouped by drug class.

Among the cephalosporins, resistance was highest for cefuroxime, with 46 (97·9%) isolates classified as resistant. Resistance was observed in 11 (23·4%) isolates for cefoperazone/sulbactam, 17 (36·2%) for ceftriaxone, and 8 (17·0%) for cefepime. Resistance to ciprofloxacin and trimethoprim/sulfamethoxazole was observed in 24 (51·1%) and 21 (44·7%) isolates, respectively. All 47 isolates were resistant to colistin.

Using the acquired-resistance categories represented in the testing panel, 18 of 47 isolates (38·3%) met the definition of multidrug resistance. Of these, one isolate was non-susceptible across all four evaluable acquired-resistance categories. MDR was more frequently observed among hospital-derived isolates, occurring in 15 of 29 (51·7%) hospital isolates compared with 3 of 18 (16·7%) community isolates, representing a significant difference between collection settings (Fisher’s exact test, OR = 5·36, p = 0·029). By subspecies, MDR was identified in 15 of 31 (48·4%) subsp. *morganii* isolates and 3 of 16 (18·8%) subsp. *intermedius* isolates, although this difference was not statistically significant (OR = 4·06, p = 0·062). A complete matrix showing the AST results for each sample has been provided in Supplementary Table S4.

### Sequence Type Diversity

The core genome phylogeny of the 47 *M. morganii* isolates, annotated by sequence type (ST), collection source (hospital or community), and β-lactamase gene profile, is shown in Figure 3. A total of 33 distinct STs were identified, demonstrating substantial sequence type diversity within the study population. Of these, 20 STs (60·6%) occurred among *M. morganii* subsp. *morganii* isolates and 13 (39·4%) among subsp. *intermedius* isolates. ST1 was the most prevalent, comprising five isolates (10·6%), followed by ST34 with four isolates (8·5%) and ST61 and ST154 with three isolates each (6·4%). All remaining STs were represented by one or two isolates. Hospital- and community-derived isolates were distributed across the phylogeny without clear source-specific clustering, and no single ST accounted for more than 10·6% of the collection.

**Figure 3:**
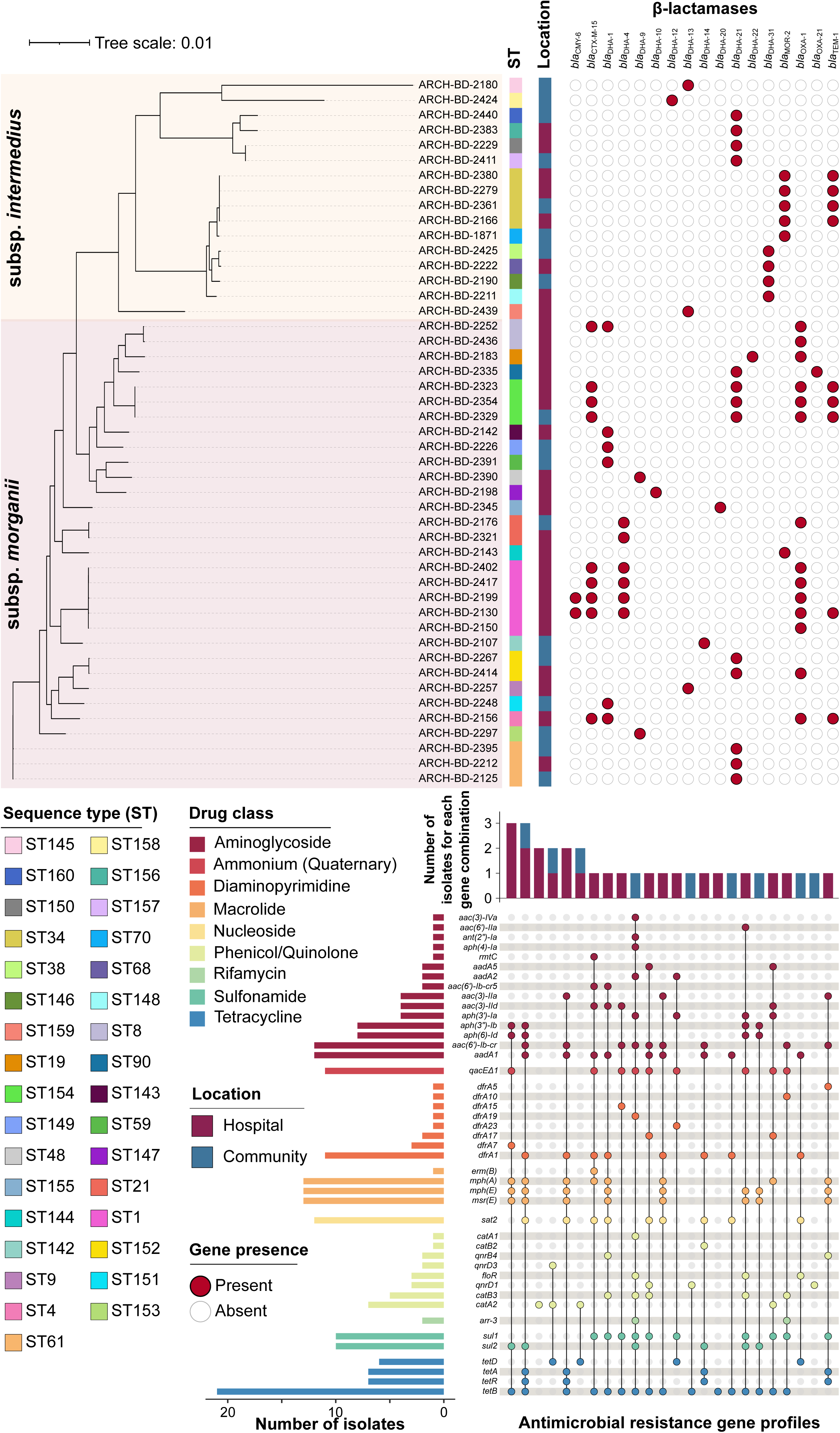
Phylogenetic relationships, sequence types, collection sources, and antimicrobial resistance gene profiles of the 47 *Morganella morganii* isolates. The core genome phylogenetic tree is annotated by sequence type (ST), collection source (hospital or community), and presence or absence of β-lactamase genes. Filled and unfilled circles indicate gene presence and absence, respectively. The UpSet plot below shows the frequency of combinations of non-β-lactam antimicrobial resistance genes across isolates. Horizontal bars indicate the total number of isolates carrying each gene, while vertical bars indicate the number of isolates carrying each gene combination. Non-β-lactam resistance genes are grouped according to the corresponding antimicrobial class.

### β-Lactamase Gene Distribution

The distribution of β-lactamase genes across the 47 isolates is also shown in Figure 3 (top) and was examined in relation to subspecies, collection source, and sequence type. A total of 17 β-lactamase alleles were identified, comprising *Morganella*-associated chromosomal AmpC-type alleles, including *bla*_MOR-2_ and multiple *bla*_DHA_ variants, together with β-lactamase determinants belonging to the CTX-M, OXA, TEM, and CMY families. The most prevalent genes were *bla*_OXA-1_ (14/47, 29·8%), *bla*_DHA-21_ (13/47, 27·7%), *bla*_CTX-M-15_ (9/47, 19·1%), and *bla*_TEM-1_ (9/47, 19·1%). Although multiple *bla*_DHA_ variants were detected, many occurred in only one or two isolates. No recognized acquired carbapenemase genes were detected among the 47 isolates.

Distinct β-lactamase profiles were observed between the two subspecies. Eleven β-lactamase genes were detected exclusively among *M. morganii* subsp. *morganii* isolates, including *bla*_OXA-1_ (14/31, 45·2%), *bla*_CTX-M-15_ (9/31, 29·0%), *bla*_DHA-1_ (6/31, 19·4%), and *bla*_DHA-4_ (6/31, 19·4%). In contrast, *bla*_DHA-12_ (1/16, 6·3%) and *bla*_DHA-31_ (4/16, 25·0%) were detected exclusively among subsp. *intermedius* isolates. Several β-lactamase determinants showed higher descriptive frequencies among hospital-derived isolates, including *bla*_CTX-M-15_ (8/29, 27·6% vs. 1/18, 5·6%), *bla*_OXA-1_ (12/29, 41·4% vs. 2/18, 11·1%), *bla*_TEM-1_ (7/29, 24·1% vs. 2/18, 11·1%), and *bla*_DHA-4_ (5/29, 17·2% vs. 1/18, 5·6%). Several STs exhibited consistent β-lactamase profiles. All four ST34 isolates carried *bla*_MOR-2_ and *bla*_TEM-1_, while all three ST154 isolates carried *bla*_CTX-M-15_, *bla*_DHA-21_, *bla*_OXA-1_, and *bla*_TEM-1_. Similarly, all five ST1 isolates carried *bla*_OXA-1_, of which four also carried *bla*_CTX-M-15_ and *bla*_DHA-4_.

### Non-β-Lactam Antimicrobial Resistance Genes

The distribution of non-β-lactam antimicrobial resistance genes across the 47 *M. morganii* isolates is shown in Figure 3 (bottom). There were 44 detected resistance genes representing nine antimicrobial or biocide resistance categories. A complete presence-absence matrix of the detected genes is provided in Supplementary Table S5.

Aminoglycoside resistance determinants represented the most diverse group, comprising 15 genes. The most prevalent were *aadA1* and the bifunctional determinant *aac(6*′*)-Ib-cr*, each detected in 12 (25·5%) isolates; *aac(6*′*)-Ib-cr* contributes to resistance to both aminoglycosides and selected fluoroquinolones. Among other frequently detected determinants, the quaternary ammonium compound resistance gene *qacE*Δ*1* and the diaminopyrimidine resistance gene *dfrA1* were each present in 11 (23·4%) isolates. The macrolide resistance genes *mph(A)*, *mph(E)*, and *msr(E)* were each identified in 13 (27·7%) isolates, while the nucleoside resistance gene *sat2* was present in 12 (25·5%). Eight genes associated with phenicol or quinolone resistance were identified but were generally less prevalent, with *catA2* being the most frequently detected at 7 (14·9%). The rifamycin resistance gene *arr-3* was detected in two isolates (4·3%). The sulfonamide resistance genes *sul1* and *sul2* were each present in 10 (21·3%) isolates. Among all non-β-lactam resistance genes identified, the tetracycline resistance determinant *tetB* was the most prevalent, occurring in 21 (44·7%) isolates.

### Single Nucleotide Variant (SNV) Analysis

Pairwise SNV matrices for *M. morganii* subsp. *morganii* and subsp. *intermedius* isolates are shown in Figure 4A and 4B, respectively. ST-specific SNV matrices, together with collection date, hospital ward, and β-lactamase profiles, are presented in Figure 4C. The SNVPhyl core genome estimates for the subspecies-level analyses were 76·47% for subsp. *morganii* and 63·27% for subsp. *intermedius*. Following stratification by ST, all core genome estimates exceeded 90%, with a minimum of 92·44%.

**Figure 4:**
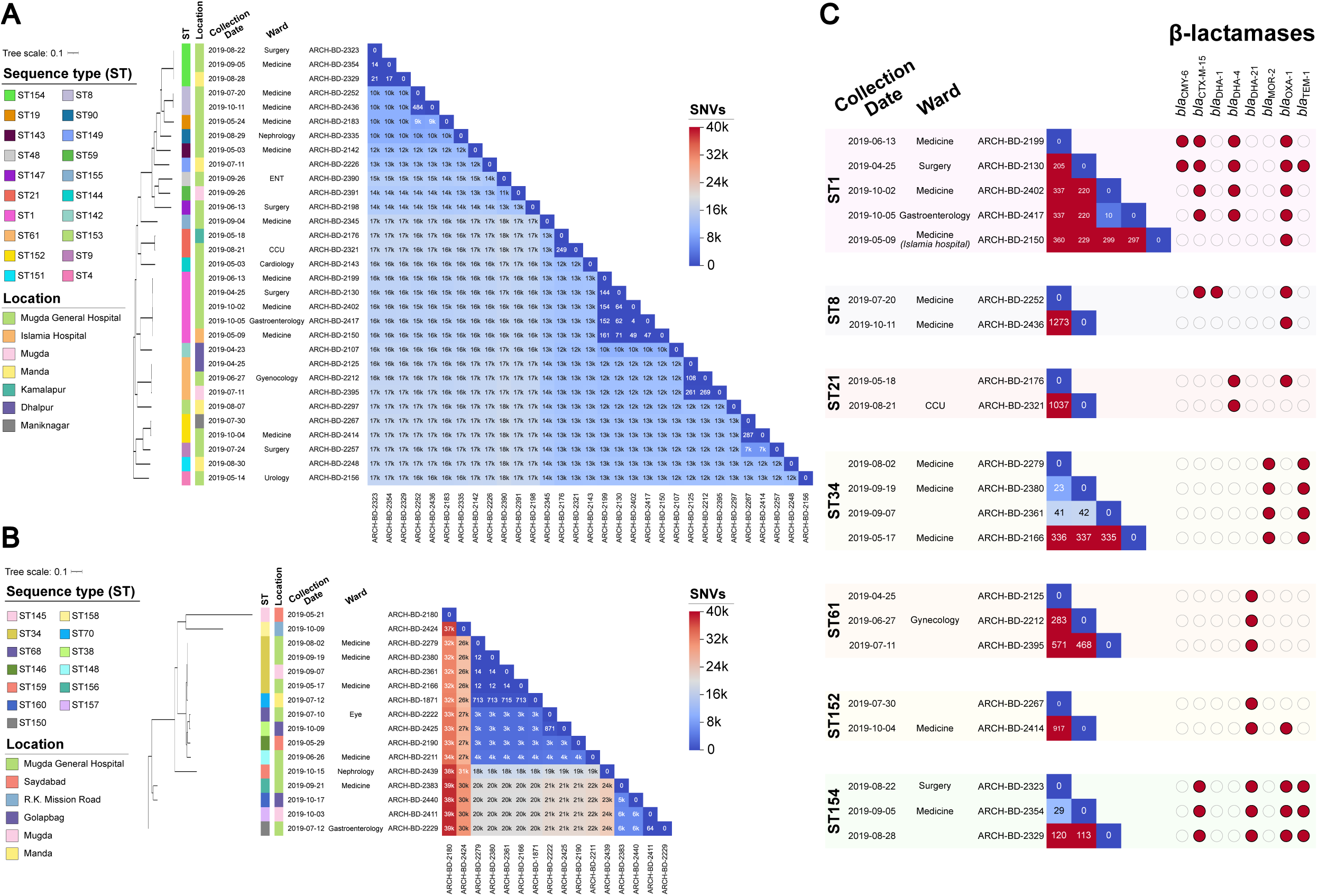
Pairwise SNV distance matrices for (A) *M. morganii* subsp. *morganii* and (B) *M. morganii* subsp. *intermedius*. Isolates are accompanied by epidemiological metadata, including sequence type, collection location, collection date, and hospital ward. (C) Pairwise SNV distance matrices for sequence types represented by multiple isolates (ST1, ST8, ST21, ST34, ST61, ST152, and ST154), shown together with collection metadata and β-lactamase gene profiles. Filled and unfilled circles indicate the presence and absence of β-lactamase genes, respectively.

Three groups showed particularly low pairwise SNV distances. Within ST1, ARCH-BD-2402 and ARCH-BD-2417 differed by 10 SNVs and were collected three days apart from different wards of the same hospital. Within ST34, ARCH-BD-2279 and ARCH-BD-2380 differed by 23 SNVs and were collected from the same hospital ward approximately seven weeks apart. A third ST34 isolate, ARCH-BD-2361, was collected from the community approximately one month after ARCH-BD-2279 and 12 days before ARCH-BD-2380, and differed from these isolates by 41 and 42 SNVs, respectively. Within ST154, ARCH-BD-2323 and ARCH-BD-2354 differed by 29 SNVs and were collected two weeks apart from different wards of the same hospital.

### Virulence-Associated Gene Distribution

The distribution of predicted virulence-associated genes across all isolates is shown in Figure 5A, with the complete presence-absence matrix provided in Supplementary Table S7. Genes were grouped according to their associated functional categories. The majority of virulence-associated genes displayed highly conserved presence/absence profiles across the study population, with little variation between isolates. The greatest variation was observed among genes associated with iron acquisition (*btuB*), oxidative stress response (*sodC*), haemolysin production (*hlyA*, *hlyB*, *hlyC*, and *hlyD*), and toxin-associated genes including toxin subunit 1 and insecticidal toxin complex components.

**Figure 5:**
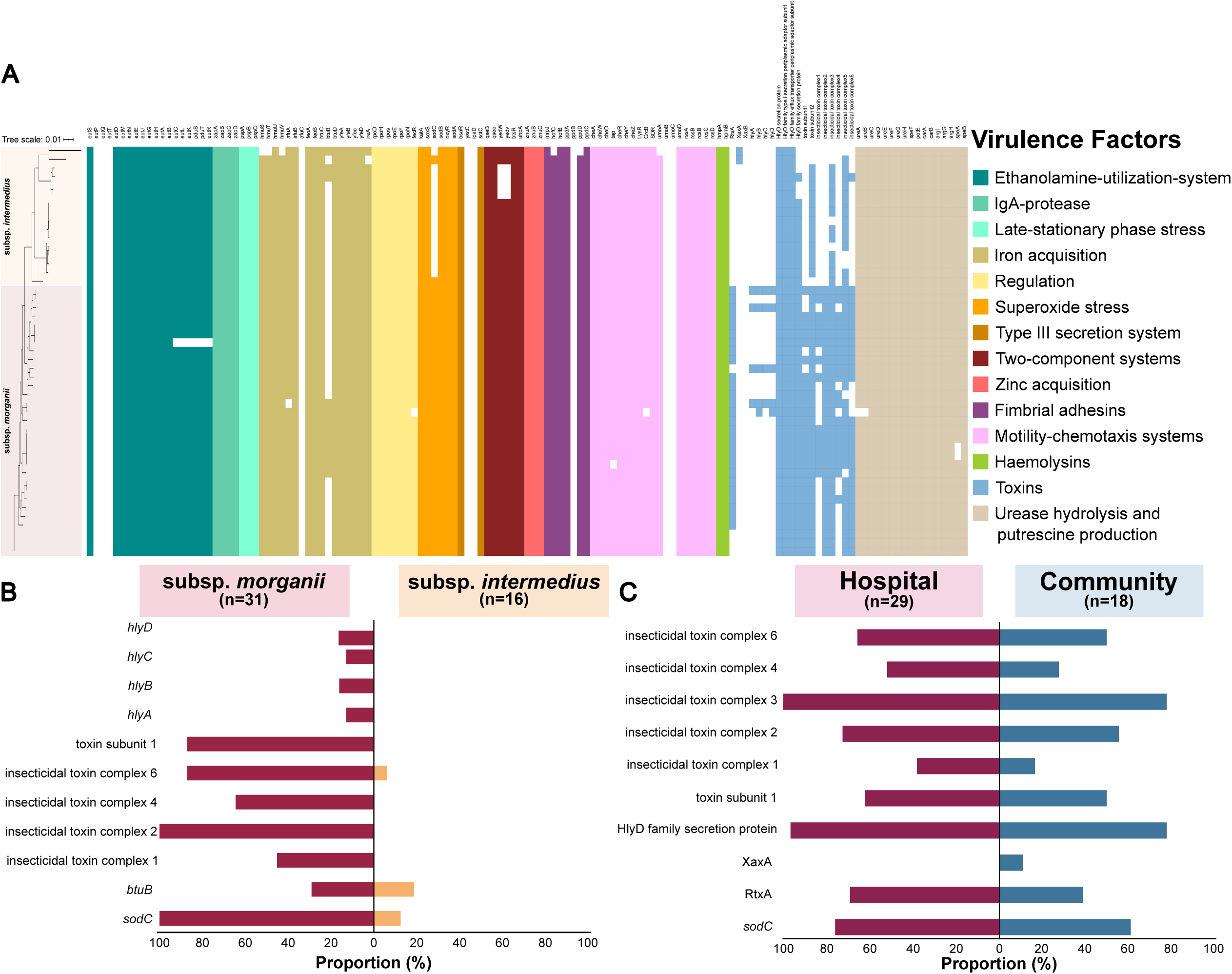
(A) Presence-absence matrix of virulence-associated genes across the 47 isolates, arranged according to the phylogenetic relationships shown in and grouped by functional category, including iron acquisition, stress response, secretion systems, adhesins, motility and chemotaxis, haemolysins, toxins, and other virulence-associated functions. (B) Comparison of the proportion of *M. morganii* subsp. *morganii* and subsp. *intermedius* isolates carrying variable virulence-associated genes (only genes differing substantially in presence between the two subspecies are shown). (C) Comparison of virulence-associated gene prevalence between hospital- and community-derived isolates (only genes showing an absolute difference in prevalence of >10 percentage points between the two groups are displayed).

The distribution of these variable virulence-associated genes between *M. morganii* subsp. *morganii* and subsp. *intermedius* isolates is presented in Figure 5B. These genes were more prevalent among the *morganii* subspecies. In contrast, subsp. *intermedius* isolates only occasionally carried these genes, with *btuB* being detected in three isolates, *sodC* in two isolates, and insecticidal toxin complex 6 detected in a single isolate.

The distribution of virulence-associated genes according to sample origin (hospital versus community) is shown in Figure 5C, which includes only genes with an absolute difference in prevalence greater than 10 percentage points between the two groups. Most variable virulence-associated genes were detected more frequently among hospital-derived isolates. The only exception was *xaxA*, which was identified exclusively in two community isolates, both belonging to the *intermedius* subspecies.

## DISCUSSION

Antimicrobial-resistant *Morganella morganii* in this Dhaka stool isolate collection comprised a diverse population spanning hospital and community settings, with no single dominant clone. Multidrug resistance was more frequent among hospital-derived isolates, while subspecies differed in genomic diversity and virulence-associated gene content. The central resistance finding was frequent imipenem resistance without detected acquired carbapenemase genes, despite uncommon resistance to meropenem and ertapenem. Together, these findings establish the value of combining phenotypic testing with genomic analysis to characterize resistance beyond carbapenemase detection alone.

Several secondary findings sharpen this interpretation. The proposed subsp. *intermedius* lineage contained greater genomic diversity and fewer hemolysin- and toxin-associated genes than subsp. *morganii*, supporting further investigation of lineage-specific biology. Two divergent genomes, including one study isolate, fell below 95% ANI relative to other genomes in these lineages, warranting taxonomic reassessment rather than a new designation based on ANI alone. Closely related isolates sampled within hospitals and across hospital and community settings identified priorities for epidemiological investigation, but did not establish direct transmission. Also of particular interest, the *aac(6*′*)-Ib-cr* gene encodes a bifunctional acetyltransferase that retains aminoglycoside-modifying activity while also reducing susceptibility to fluoroquinolones such as ciprofloxacin and norfloxacin, illustrating how individual acquired determinants may contribute to resistance across multiple antimicrobial classes.^45^ The coexistence of intrinsic and acquired resistance determinants also underscores the need to distinguish species-characteristic resistance from acquired multidrug resistance.

These findings extend the phenotypic ARCH surveillance in Bangladesh by adding genomic resolution to resistance observed across hospital and community settings.^46^ The population structure agrees with Bonnin and colleagues’ genomic framework,^12^ while the diversity within subsp. *intermedius* complements subsequent work treating it as a distinct genospecies whose taxonomic rank remains unsettled.^47^ Differences in virulence gene content are consistent with previously reported variation among *M. morganii* lineages.^11^ The imipenem phenotype parallels Zheng and colleagues’ carbapenemase-negative isolates, in which reduced *lpoA*/*lpoB* expression, penicillin-binding protein mutations, and efflux activity were implicated.^8^ However, carbapenemase genes have also been reported in isolates with selective imipenem resistance;^48^ the phenotype therefore does not identify a single mechanism. These published mechanisms remain hypotheses for our isolates.

The study’s principal limitation is selection of antimicrobial-resistant isolates from one urban setting: observed resistance frequencies cannot estimate prevalence among all *M. morganii* in Dhaka or Bangladesh. The small sample limited comparisons, and differences between collection settings could reflect lineage composition or other unmeasured factors; antibiotic exposure and hospital acquisition were not established as causes. Low core genome coverage constrained subspecies-wide SNV comparisons, requiring analysis by sequence type. Cross-sectional sampling, limited epidemiological metadata, and the absence of a validated transmission threshold prevented reconstruction of transmission chains. Short reads limited resolution of resistance-gene location and mobility, and the susceptibility panel did not support XDR or PDR classification. Gene detection did not establish expression, resistance mechanisms, or phenotypic virulence. Broader sampling and functional data could therefore change estimates of resistance burden and interpretations of lineage differences.

These results support including *M. morganii* in genomic AMR surveillance alongside phenotypic susceptibility testing. Larger longitudinal studies should sample both resistant and susceptible isolates from carriage, clinical infections, and potential reservoirs, with detailed exposure and contact data. Long-read sequencing and functional assays should resolve resistance-gene context, test the basis of imipenem resistance, and determine whether lineage-associated gene differences affect virulence. Linking these genomic patterns to mechanisms and routes of spread is the next step toward identifying where prevention can interrupt the circulation of resistant *M. morganii*.

## CONCLUSION

This study demonstrates that stool-derived *M. morganii* circulating among community and hospital populations in Dhaka comprises a genetically diverse population with distinct subspecies, extensive antimicrobial resistance, and subspecies-associated differences in virulence gene content. Frequent imipenem non-susceptibility in the absence of recognized acquired carbapenemases highlights the need to distinguish species-characteristic reduced susceptibility from acquired chromosomal or regulatory resistance mechanisms that may not be captured through conventional AMR gene screening alone. Several closely related isolates were also identified across hospital wards and between community and hospital settings, although the available genomic and epidemiological data were insufficient to establish direct transmission. Collectively, these findings support greater consideration of *M. morganii* in genomic AMR surveillance in Bangladesh and highlight the need for broader longitudinal studies to define its population structure, resistance mechanisms, ecological reservoirs, and potential transmission pathways.

## ACKNOWLEDGMENTS

We gratefully acknowledge icddr,b core donors (Govt. of Bangladesh and Canada) for their unrestricted support and commitment to icddr,b’s research efforts. The study was funded by the Centers for Disease Control and Prevention, Atlanta, GA, USA, through a cooperative agreement. We also thank Dr. Kara Moser (Clinical and Environmental Microbiology Branch, Division of Healthcare Quality and Promotion, Centers for Disease Control and Prevention, Atlanta, GA, USA) for her advice and suggestions regarding the methodology.

## Ethical considerations

The protocol was reviewed and approved by the Research Review Committee and Ethical Review Committee of icddr,b (protocol PR18060). Written informed consent was obtained from participating human subjects according to the approved protocol. All patient IDs were de-identified to isolate IDs which were not known to anyone outside the research group, and only deidentified isolate IDs are provided in the manuscript.

## Transparency statement

The corresponding author affirms that this manuscript is an honest, accurate, and transparent account of the study being reported; that no important aspects of the study have been omitted; and that any discrepancies from the study as planned (and, if relevant, registered) have been explained.

## Data Availability

The whole-genome sequencing data generated in this study have been deposited in the NCBI Sequence Read Archive (SRA) under BioProject accession PRJNA962904. The individual SRA accession numbers for all isolates included in this study are provided in Supplementary Table S2.

